# Validation of breath biomarkers for obstructive sleep apnea

**DOI:** 10.1101/2021.03.16.21253612

**Authors:** Nora Nowak, Anna Engler, Sira Thiel, Anna S. Stöberl, Pablo Sinues, Renato Zenobi, Malcolm Kohler

## Abstract

**Background and objectives:** Obstructive sleep apnea (OSA) is an underdiagnosed respiratory disease with negative metabolic and cardiovascular effects. The current gold standard for diagnosing OSA is in-hospital polysomnography, a time-consuming and costly procedure, often inconvenient for the patient. Recent studies revealed evidence for the potential of breath analysis for the diagnosis of OSA based on a disease-specific metabolic pattern. However, none of these findings were validated in a larger and broader cohort, an essential step for its application in clinics.

**Methods:** In the present study, we validated a panel of breath biomarkers in a cohort of patients with possible OSA (N = 149). These markers were previously identified in our group by secondary electrospray ionization high-resolution mass spectrometry (SESI-HRMS).

**Results:** Here, we could confirm significant differences between metabolic patterns in exhaled breath from OSA patients compared to control subjects without OSA as well as the association of breath biomarker levels with disease severity. Our prediction of the diagnosis for the patients from this completely independent validation study using a classification model trained on the data from the previous study resulted in an area under the receiver operating characteristic curve of 0.66, which is comparable to questionnaire-based OSA screenings.

**Conclusions:** Thus, our results suggest that breath analysis by SESI-HRMS could be used to screen for OSA. Its true predictive power should be tested in combination with OSA screening questionnaires.

**Clinical trial:** **“**Mass Spectral Fingerprinting in Obstructive Sleep Apnoea”, NCT02810158, www.ClinicalTrials.gov

## 1. Introduction

Obstructive sleep apnea (OSA) is a highly prevalent sleep-related breathing disorder.[1] The repeated partial or complete collapse of the pharynx during sleep provokes apnea or hypopnea events, which may lead to repetitive oxygen desaturations. Frequent sleep disruptions and increased activity of the sympathetic nervous system are accompanying these apnea/hypopnea events resulting in poor sleep quality and increased daytime sleepiness.[2] Several metabolic and cardiovascular consequences, such as an increased risk for cardiovascular disease, arterial hypertension, diabetes, vascular dysfunction, as well as depression, are well known.[3–6] OSA can be effectively treated i.e. with continuous positive airway pressure (CPAP).[7–9]

The conventional diagnosis of OSA is carried out by respiratory polygraphy or even polysomnography, [10,11] which are time-consuming, costly and inconvenient for patients. In addition, there is emerging evidence for a high night-to-night variability of OSA, posing another challenge for diagnostics.[12] Thus, for a reliable diagnosis, testing during several nights would be required. Screening for OSA is conventionally based on questionnaires, such as the Epworth Sleepiness Scale,[13] the STOP-bang,[14] Berlin[15] or NoSAS score.[16] However, the results from such questionnaires are by nature subjective.

Exhaled breath contains several hundreds of metabolites and thus provides insights into biochemical processes of the human body.[17] Many of the metabolites in breath do not originate from the lungs but are transported from blood to the airways via gas exchange in the lung. Therefore, breath metabolite levels mostly reflect systemic metabolic processes. Furthermore, consistent alterations of the molecular fingerprint of exhaled breath in patients with a certain disease may indicate disease specific metabolic changes. Such disease specific biomarkers detected in exhaled breath could be the basis for an objective and non-invasive diagnostic procedure, which is fast and easy to perform for patients.

So far, many studies with small sample sizes have obtained promising results, suggesting a great diagnostic potential of exhaled breath analysis for various diseases. However, larger validation studies are missing and, to date, exhaled breath analysis is applied in clinical routine only for very few applications, such as the evaluation of bronchial inflammation by measuring fractional exhaled nitric oxide (FeNO).[18] To achieve a more widespread clinical application of breath analysis for disease diagnosis and monitoring, the validation of preliminary findings in large cohorts of patients is essential.

The investigation of exhaled breath in OSA patients using different technical approaches revealed convincing results for diagnosing this disease and for monitoring patients’ compliance to CPAP therapy.[19] In some studies, electronic sensors (e-noses) were used to recognize OSA specific patterns in exhaled breath.[20–23] In their attempts of diagnosing OSA against the gold-standard (polysomnography), areas under the receiver operating characteristic curves (AUROCs) in the range from 0.84 (no 95% CI provided) to 0.87 (95% CI 0.61−1.00) were reported, suggesting future diagnostic applicability of breath analysis. However, e-noses do not allow for compound identification and thus do not provide mechanistic insights into the disease, but merely produce a complex “signal” whose statistical evaluation can give some valuable output. Furthermore, the data from studies with e-noses were not validated in larger and broader cohorts of patients with possible OSA.

An untargeted investigation from our laboratories of an extensive spectrum of molecules in exhaled breath using secondary electrospray ionization high-resolution mass spectrometry (SESI-HRMS) revealed specific markers, allowing identification of a disease specific molecular profile of exhaled breath in patients with OSA recurrence after two weeks of CPAP therapy withdrawal.[24] In that randomized controlled trial, significant correlations between metabolite levels in breath and change in oxygen desaturation index (ODI) upon CPAP withdrawal, and significant differences between the CPAP withdrawal and treatment group were found. Further, we achieved a successful classification (AUROC = 0.87) between the withdrawal group and the group that continued the treatment. In order to transfer our promising findings into the diagnostic algorithm of OSA, this observational study aims to validate these metabolic breath profiles in a larger treatment naive cohort of patients with possible OSA.

## 2. Methods

### 2.1 Study participants

This study includes 149 participants with possible OSA in the age of 53.3 ± 13.7 years with a BMI of 30.1 ± 6.6 kg/m^2^ (table 1). The study protocol was approved by the local ethical committee (KEK-ZH 2016-00384). The experiments were conducted in accordance with the Declaration of Helsinki and written informed consent was obtained from all participants before participation. The clinical trial was registered at ClinicalTrials.gov (NCT02810158).

**Table 1:**
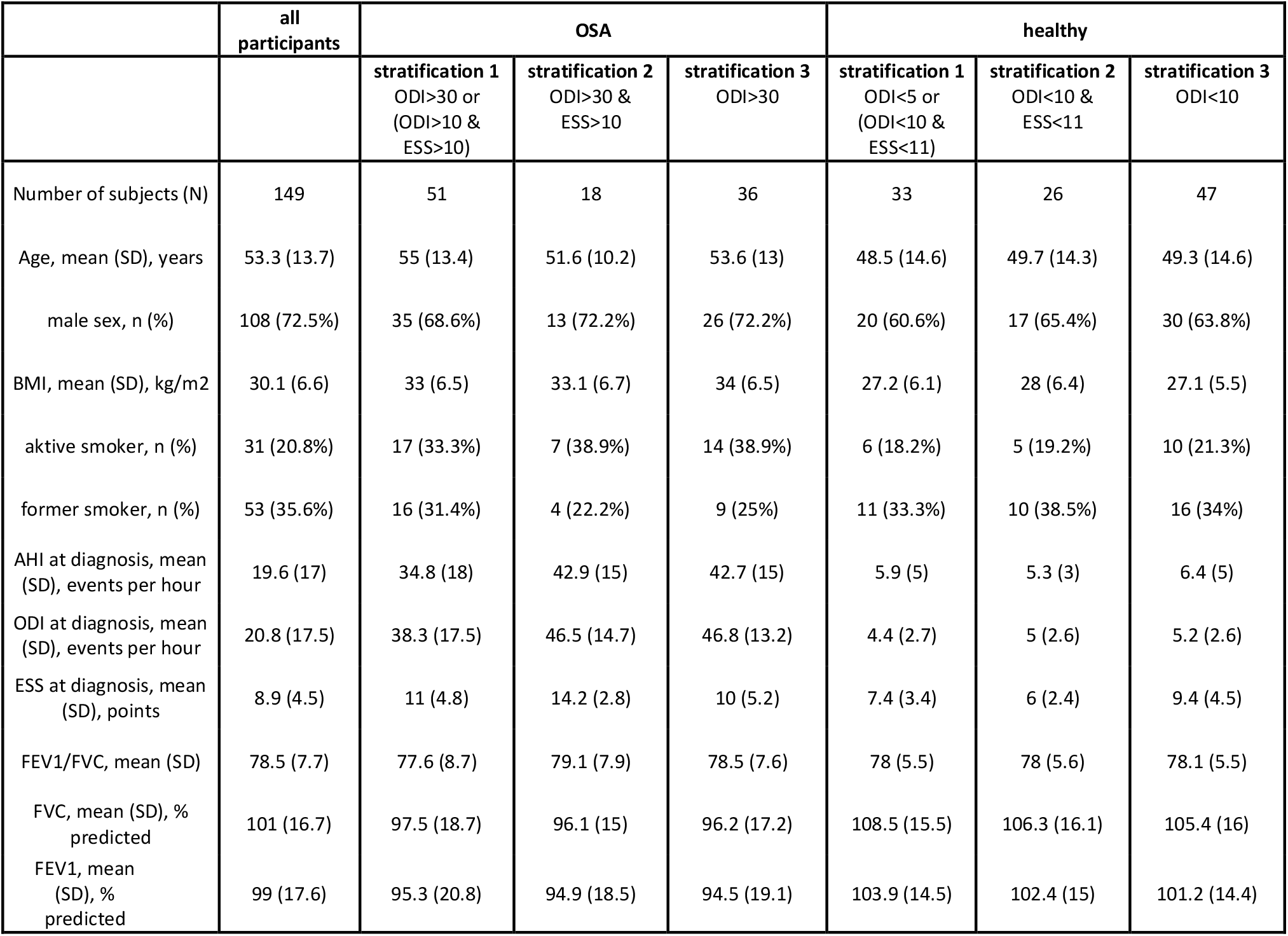
Study participant characteristics. (AHI: apnea-hypopnea-index; BMI: body mass index; ESS: Epworth Sleepiness Scale; ODI: oxygen desaturation index; FVC: expiratory forced vital capacity; FEV1: forced expiratory volume in one second)

All patients underwent in-hospital respiratory polygraphy (RP). Inpatient RPs were recorded by Alice 6 Diagnostic System (Philips Respironics, PA, USA), scored with validated Somnolzyer 24×7 software (Philips Respironics, PA, USA), and reviewed manually. The data obtained was evaluated according to the guidelines of the American Academy of Sleep Medicine.[13] The participants were asked to fill in Epworth Sleepiness Scale (ESS) questionnaires.

### 2.2 SESI-HRMS measurements

Participants were asked to refrain from eating, drinking, chewing gum, alcohol, tobacco, caffeine use or brushing their teeth at least 1 hour prior to the SESI-HRMS measurements. Exhaled breath of 149 patients was analyzed by SESI-HRMS using a commercial SESI source (SEADM, Spain) coupled to a high-resolution TripleTOF 5600+ mass spectrometer (AB Sciex, Concord, Ontario, Canada). The participants were sitting in upright position in front of the mass spectrometer and exhaled at least six times with a pressure drop of 12 mbar through a disposable mouthpiece into the heated sampling line, connected to the SESI source. A flow splitter at the front-end enabled sampling of end-tidal breath. The flow through the ion source was set to 0.2 L/min. Full scan mass spectra were recorded in positive ion mode with an accumulation time of 1 s in the rage of 50-500 Da.

### 2.3 Data preprocessing

All mass spectral data was analyzed with MATLAB R2020a and R 4.0.0. Mass spectra obtained from exhaled breath were preprocessed as described elsewhere.[25] In short, mass spectra were interpolated, aligned, exhalation time windows were chosen and peak picking was performed on the average breath spectrum. As in the pilot study[24], breath signal intensities were normalized to the median intensity of the total ion current and then autoscaled.

### 2.4 Statistical analysis

Further, the features were filtered for markers that have been associated with OSA previously[24]. The mass-to-charge ratio (m/z) tolerance was set to 0.005 Da. The remaining 78 m/z features were first tested for normality in a Shapiro-Wilk’s test. Since the data was not normally distributed (p-value distribution from Shapiro-Wilk’s test for normality is provided in Figure A1), we performed a correlation analysis between signal intensity and ODI as well as between signal intensity and ESS using Spearman correlation.

Moreover, we tested for differences in signal intensities between individuals without OSA and OSA patients by performing two-sided Mann-Whitney-U tests. Here, we first applied stratification criteria as they are commonly applied in the clinics: OSA ODI > 30/h or ODI > 10/h & ESS > 10 points, control ODI < 5/h or ODI < 10/h & ESS < 11 points (stratification 1). We then also tested for between-group differences with stricter stratification criteria (OSA: ODI > 30/h & ESS > 10 points, control: ODI < 10/h & ESS < 11 points, stratification 2) in order to remove individuals with ambiguous diagnosis. We also calculated log_2_ fold changes between the groups. To account for multiple hypothesis testing, false discovery rates (q-values) were calculated for all obtained p-values using Storey’s procedure.[26]

### 2.5 Classification Procedure

We combined the breath intensities obtained for the 78 m/z features mentioned above of the previously reported pilot study and this validation study. We used the data of the pilot study as training set and used the MATLAB classification learner app to find the best classification algorithm. For model evaluation we used a 7-fold cross validation. We defined the OSA and control group with the above mentioned criteria of stratification 1. In order to obtain balanced group sizes we only used the before and after measurements of the 9 individuals of the placebo group, who developed OSA in the previous study.[24] A Gaussian support vector machine model performed best. We thus trained such model on the training data and predicted the validation data set obtained from this study.

### 2.6 Attempts of improvement of classification performance

Since both data sets were acquired on different mass spectrometers and with different generations of SESI sources, we assessed by principal component analysis (PCA) the comparability of both data sets. A slight shift between both data sets was observed. We therefore performed a batch correction based on an empirical Bayes algorithm[27] and repeated the classification procedure described above (Figure A2). We also repeated the classification procedure with stratification criteria, which are more similar to the ones used in the pilot study. For the validation data set we defined the groups as follows: OSA: ODI > 30/h, control: ODI < 10/h. In order to get balanced group sizes in the training set, we reduced the control group to ODI < 2/h.

## 3. Results

### 3.1 Study design and patient characteristics

149 participants between 19 and 83 years with possible OSA underwent respiratory polygraphy and breath analysis by SESI-HRMS directly after the sleep study (Figure 1). Patient characteristics and results from respiratory polygraphy are shown in Table 1. Depending on the applied stratification criteria, the mean ODI in the OSA group varies between 38.3 and 46.8 events per hour. The control group had a mean ODI between 4.4 and 5.2 events per hour. The mean Epworth Sleepiness Scale score (obtained from a questionnaire estimating the extend of daytime sleepiness) in the OSA group ranged from 10 to 11 points and in the control group from 6 to 9.4 points.

**Figure 1:**
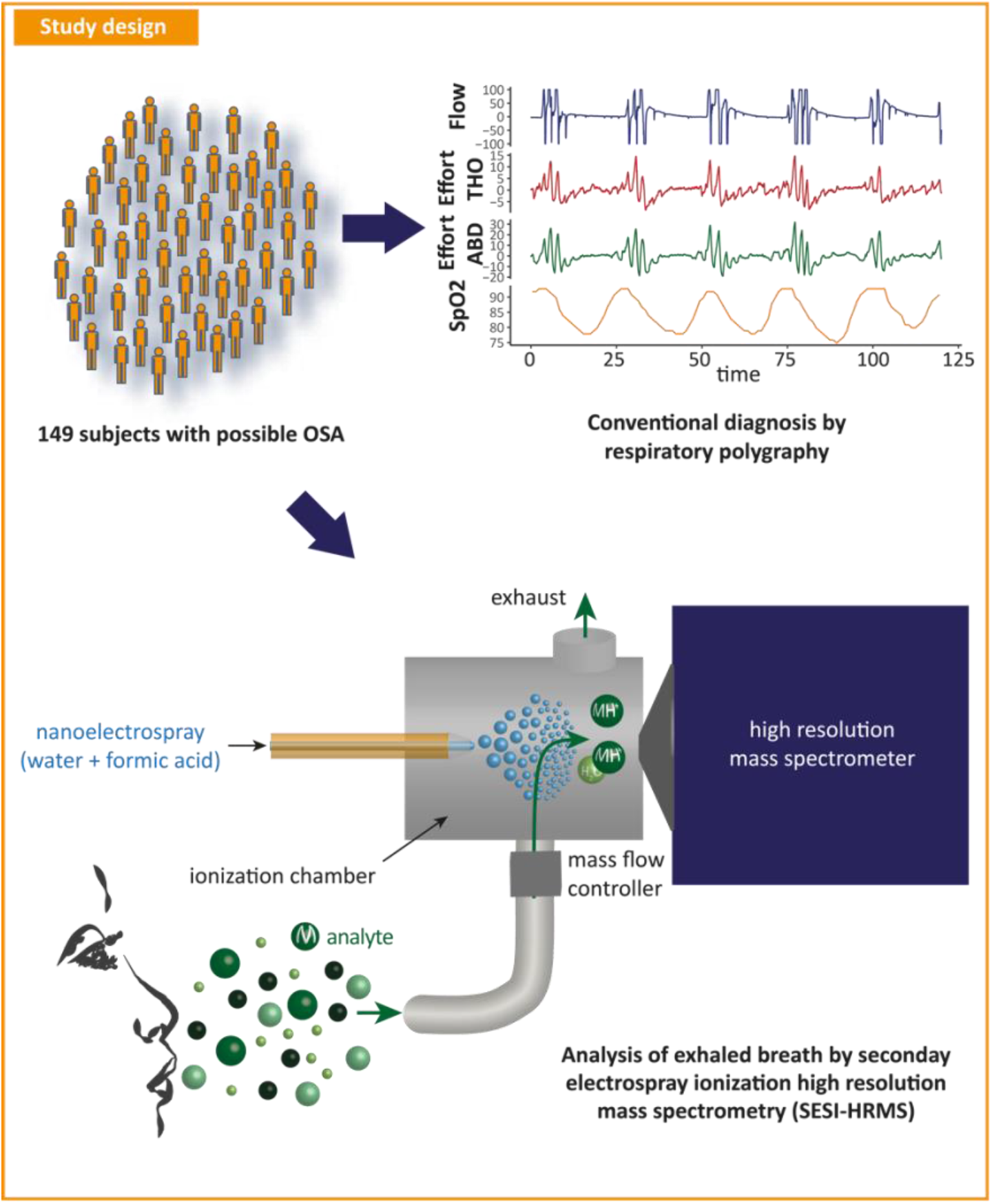
Study design. 149 study subjects with possible OSA underwent conventional diagnosis by respiratory polygraphy in the sleep laboratory and exhaled breath analysis by SESI-HRMS.

### 3.2 Metabolic patterns in exhaled breath associated with OSA

The data obtained in this validation study from SESI-HRMS measurements was pre-processed in the same way as it was done in our previous study[24], which we refer to as pilot study in the following, i.e., the signal intensities were normalized to the median of total ion current and then autoscaled. We continued our analysis in a targeted fashion focusing only on the m/z features that have previously been associated with OSA in our pilot study. However, we were not able to detect all of them, which is most likely due to technical optimization of the acquisition method that was made in the meantime. Nevertheless, 78 of the features that have been previously reported either as significantly different between the CPAP and the withdrawal group, or as correlating with the change in ODI or as predictive for OSA, were also detected in this validation study. For those 78 m/z features, we tested for significant differences between controls (without OSA) and OSA patients and for correlation with ODI and ESS. Moreover, we trained a classification model with the data from the pilot study and predicted the OSA diagnosis of the validation cohort from this study.

### 3.3 Significant differences in metabolic breath patterns between OSA patients and individuals without OSA

We tested for significant differences in metabolite intensities in exhaled breath between OSA patients and controls without OSA (Mann-Whitney-U test). We assigned the participants to two groups (OSA and control) based on the following criteria, commonly applied in clinics: ODI > 30/h or ODI > 10/h & ESS > 10 points (definitive OSA); and ODI < 5/h or ODI < 10/h & ESS < 11 points (definitive without relevant OSA; control) (stratification 1). All subjects in between were assigned to an “unclear” group, since no unambiguous OSA diagnosis could be stated. For 19 features we found significant (p ≤ 0.05) differences between the two groups (Figure 2A-C, boxplots of two examples are shown in Figure 2D and E, all boxplots are provided in Figure A3). When we used stricter grouping criteria (OSA: ODI > 30/h & ESS > 10 points, control: ODI < 10/h & ESS <11 points, stratification 2) in order to consider only patients with an unambiguous diagnosis, significance increases as shown in Figure A4 (all boxplots are given in Figure A5). All numeric results are provided in Table A1. Hence, our results from this validation study confirm our previous findings of a specific metabolic pattern in exhaled breath in OSA patients.

**Figure 2:**
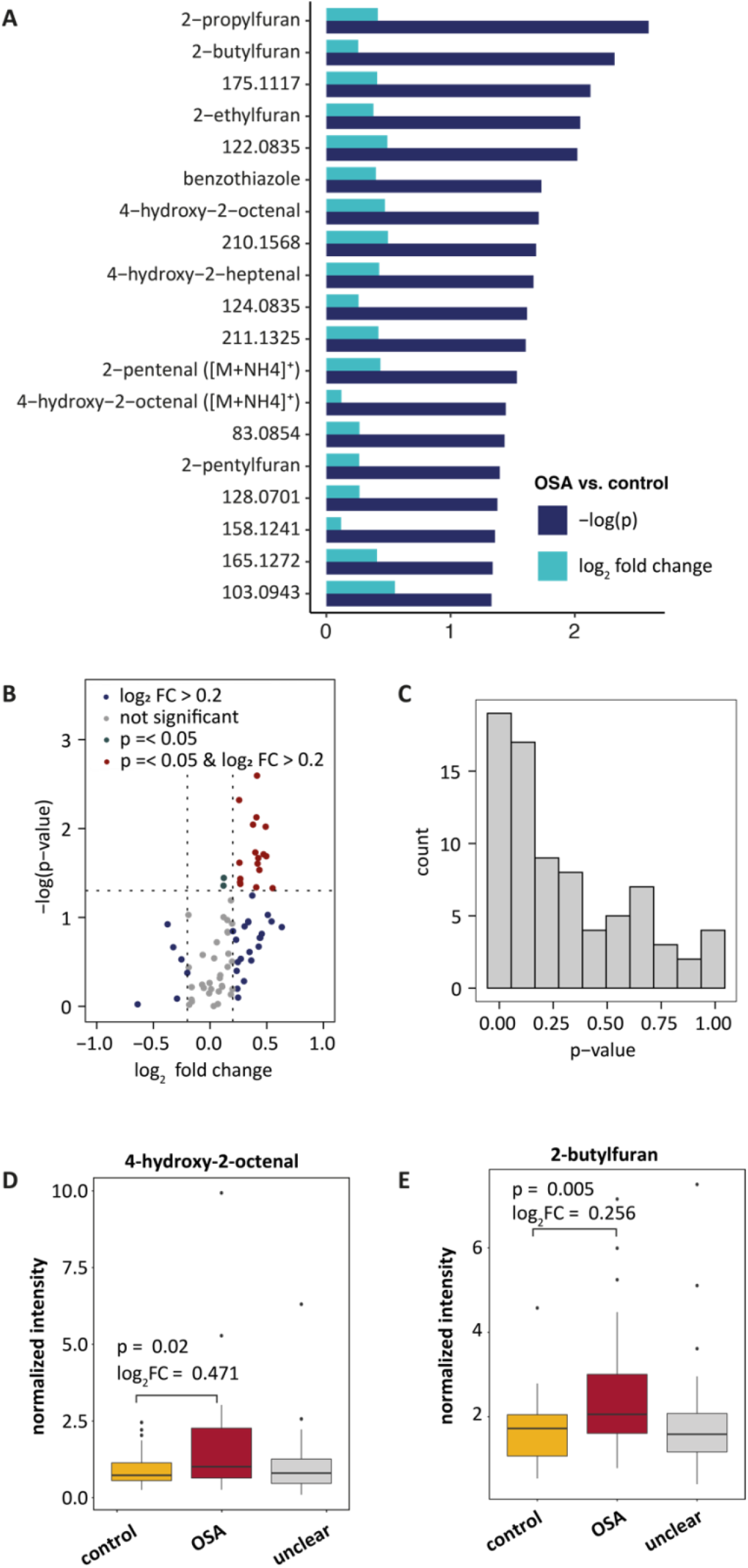
Significant differences in metabolic breath patterns between subjects with and without OSA. (OSA: ODI > 30/h or ODI > 10/h & ESS > 10 points; control: ODI < 5/h or ODI < 10/h & ESS < 11 points; unclear: in between; stratification 1). **A** p-values and fold changes of significant features sorted by significance. Not identified features are labelled with their m/z. **B** volcano plot for all 78 metabolites. **C** p-value distribution for between-group differences from Mann-Whitney-U test. **D, E** Exemplary boxplots (center line: median, box limits: 25th and 75th percent quantile, whisker length: 1.5 interquartile range) of 4-hydroxy-2-octenal and 2-butylfuran. The Boxplots for all significant features are provided in Figure A1, numeric results for significant features are summarized in Table 2 and numeric results of all 78 features are given in Table A1.

### 3.4 Association between disease severity and breath signal intensity

To test whether the breath patterns are also correlating with the severity of OSA in this larger and more diverse cohort of patients, we performed a Spearman correlation analysis. For 21 features, we found a significant correlation between breath levels and ODI (p ≤ 0.05). (Figure 3A, correlation plots for two examples are shown in Figure 3B, all correlation plots are provided in Figure A6) All except one show higher intensities for an increased ODI, suggesting that oxygen desaturation correlates with an enrichment of these metabolites. Amongst these metabolites correlating with the ODI are several unsaturated aldehydes as well as furanes and benzothiazole that have been identified before.[28–30] Thus, we could confirm the previously reported association between disease severity and breath signal intensity.

**Figure 3:**
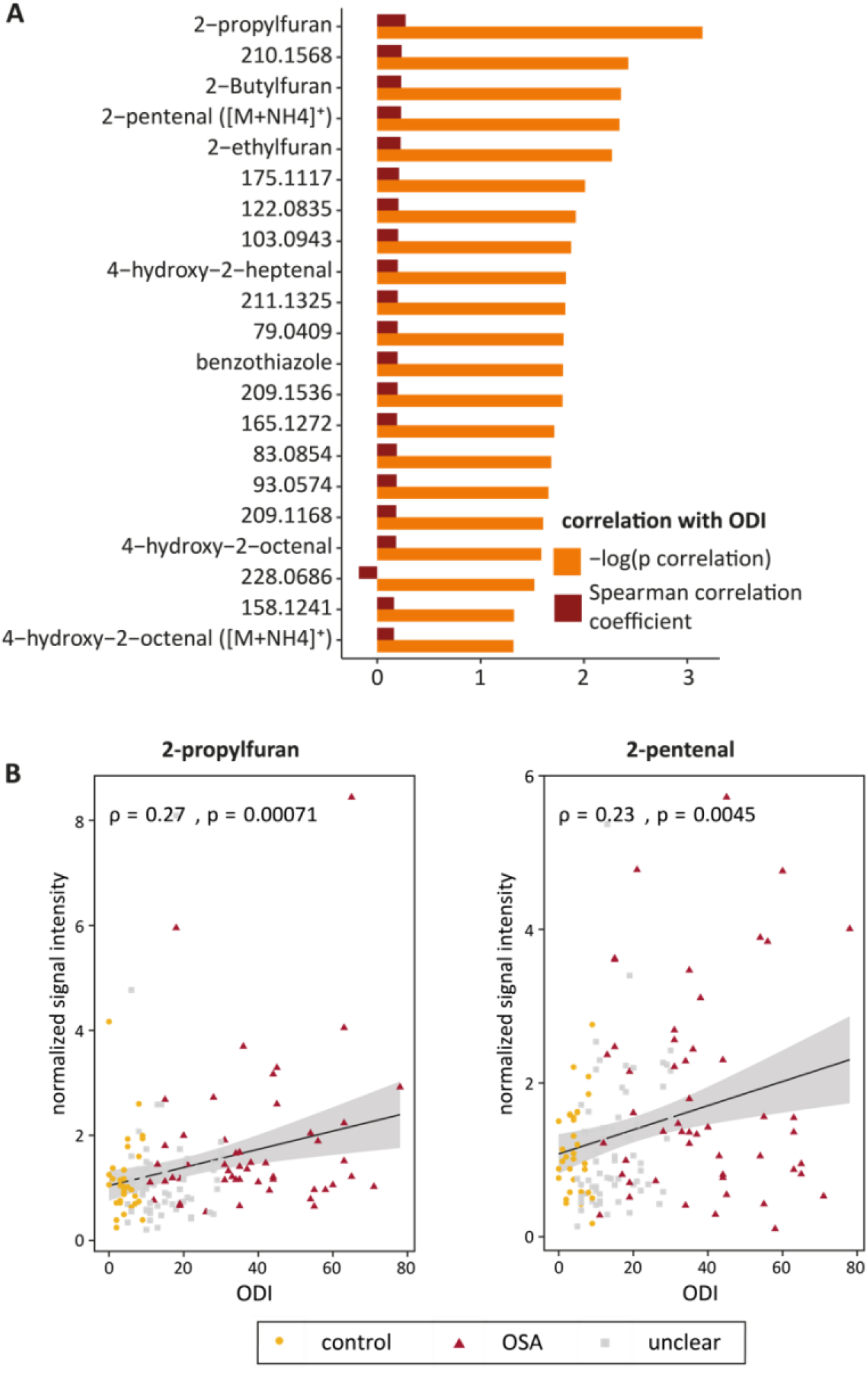
Correlations between metabolite levels in breath and OSA severity. **A** p-values and correlation coefficients of features from exhaled breath with significant correlations with the ODI. Not identified features are labelled with their m/z. **B** Exemplary regression lines for 2-propylfuran and 2-pentenal. Regression lines for all features with significant correlations with the ODI are provided in Figure A4. Numeric results for features with significant correlations are summarized in Table 2 and numeric results of all 78 features are given in Table A1. (OSA: ODI > 30/h or ODI > 10/h & ESS > 10 points; control: ODI < 5/h or ODI < 10/h & ESS < 11 points; unclear: in between; stratification 1)

### 3.5 Association between sleepiness and breath signal intensity

For nine features we found a significant correlation between their breath intensities and the ESS (p ≤ 0.05) (Figure 4A). Amongst them, four features furthermore correlate with ODI, such as 2-pentylfuran and 4-hydroxy-2-octenal. Correlation plots for these two examples are shown in Figure 4B (all correlation plots are provided in Figure A7).

**Figure 4:**
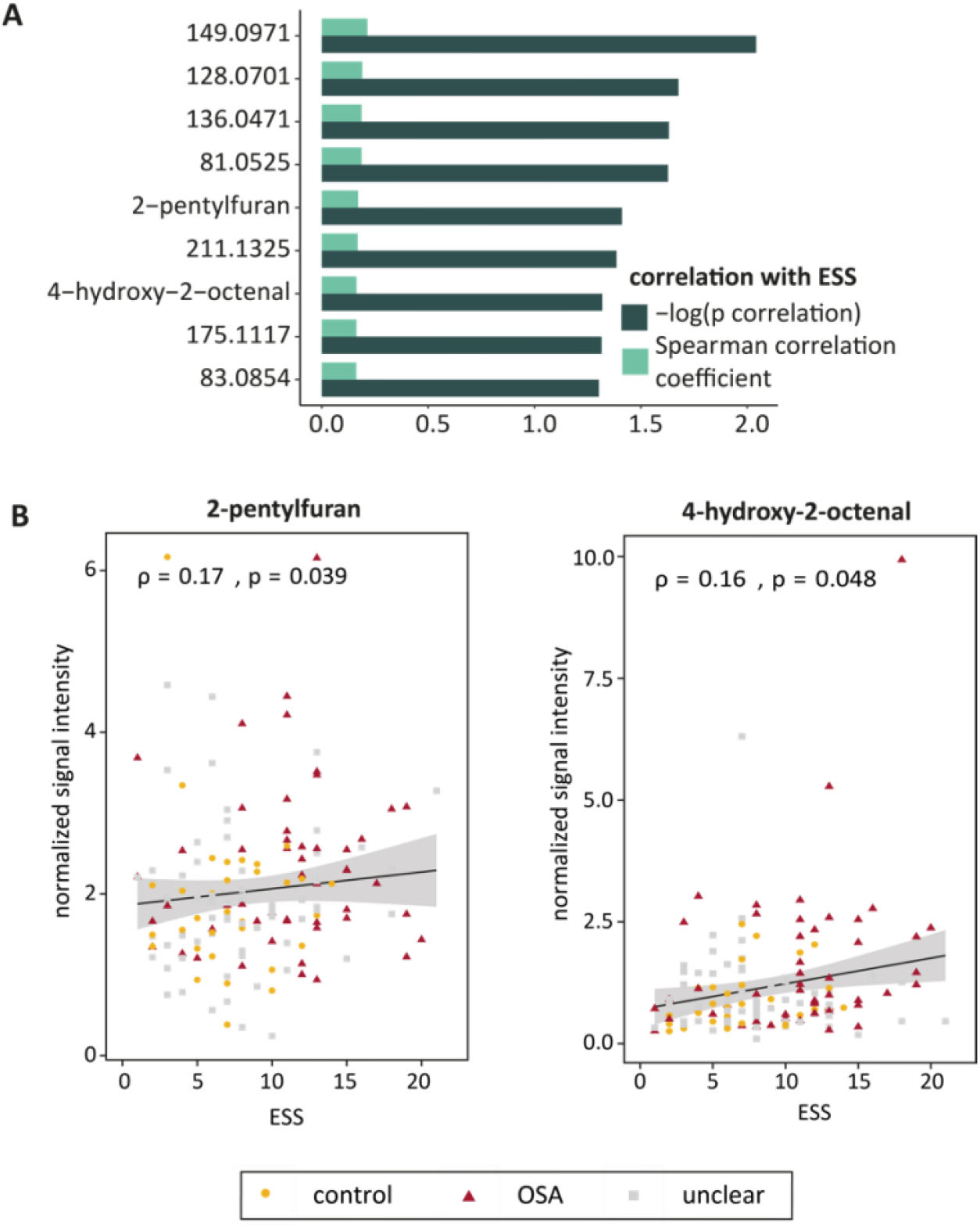
Correlations between metabolite levels in breath and sleepiness. **A** p-values and correlation coefficients of features from exhaled breath with significant correlations with the ESS. Not identified features are labelled with their m/z. **B** Exemplary regression lines for 2-pentylfuran and 4-hydroxy-2-octenal. Regression lines for all features with significant correlations with the ESS are provided in Figure A5. Numeric results for features with significant correlations are summarized in Table 2 and numeric results of all 78 features are given in Table A1. (OSA: ODI > 30/h or ODI > 10/h & ESS > 10 points; control: ODI < 5/h or ODI < 10/h & ESS < 11 points; unclear: in between; stratification 1)

### 3.6 Classification

To assess to applicability of metabolite levels measured in exhaled breath using SESI-HRMS for the clinical diagnosis of OSA, we trained a classification model with the data from our pilot study and predicted the diagnosis of OSA or control in the validation cohort measured in this study. Figure 5 shows the classification procedure schematically.

**Figure 5:**
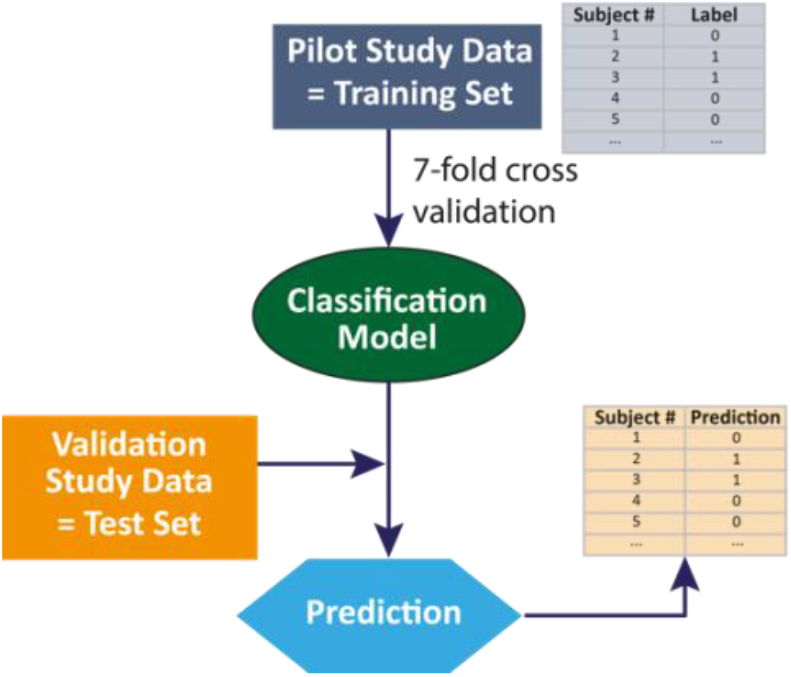
Classification procedure. A classification model was trained with the data from our pilot study and its performance was estimated in a 7-fold cross validation. Subsequently, the diagnosis of the patient cohort from this validation study was predicted.

First, we grouped the patients again as described above by the clinical criteria of stratification 1. In order to obtain balanced group sizes in the training set, we used only the “before” and “after” measurements of those patients in the CPAP withdrawal group, who developed significant OSA under placebo treatment (Figure 6A). With the training data, we estimated the performance of the classification model in a 7-fold cross-validation. This resulted in an AUROC of 0.59 (Figure 6B, the confusion matrix is provided in Figure 6C). The prediction of the diagnosis for the validation data (Figure 6D) yielded in an AUROC of 0.66 (Figure 6B, confusion matrix is given in Figure 6C). The accuracy of the prediction was 63% with a sensitivity of 76% and a specificity of 42%.

**Figure 6:**
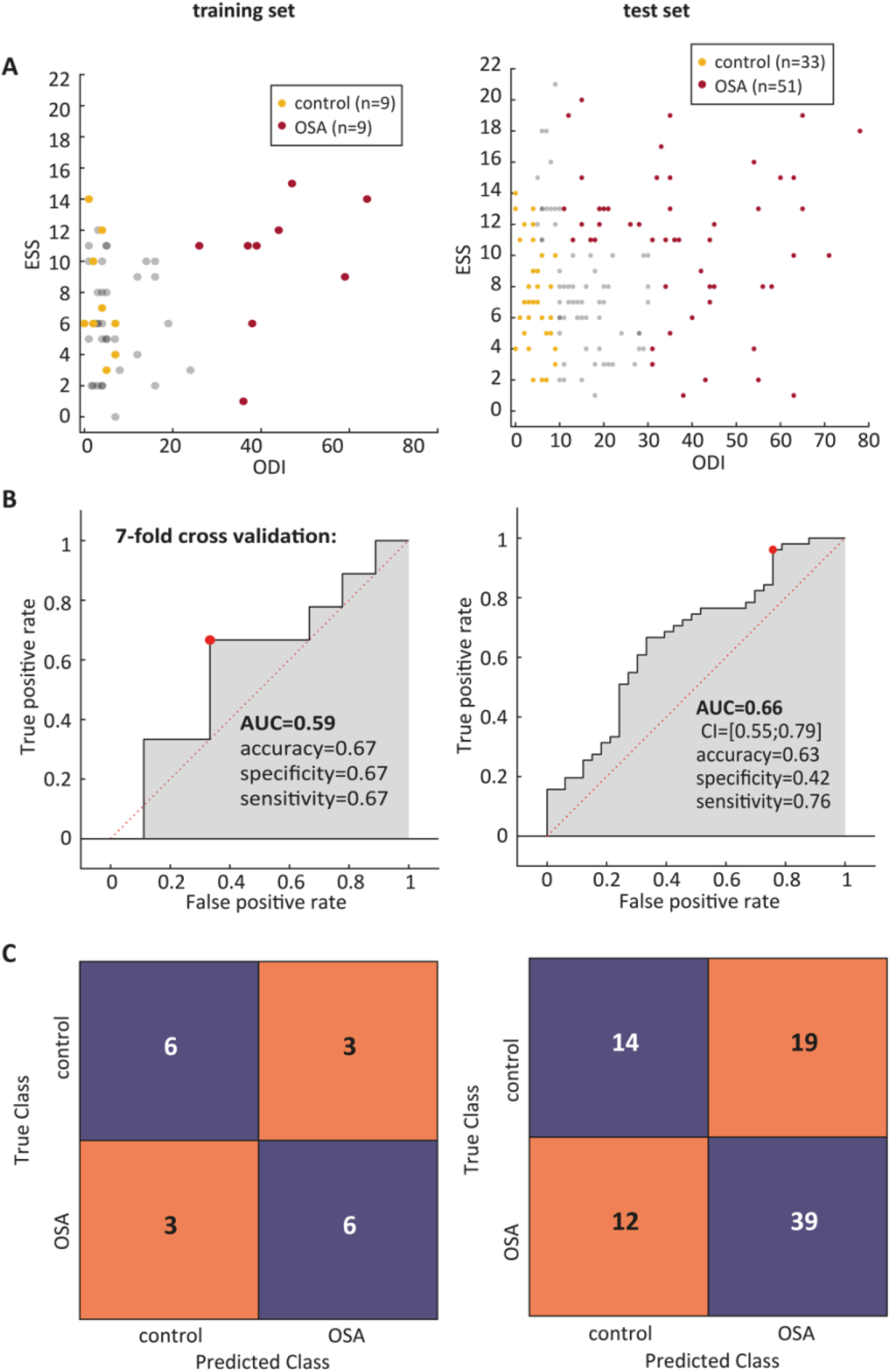
Classification results. **A** ESS and ODI of samples in the training set (left) and test set (right). **B** ROC curve from 7-fold cross validation of the classification model with the training set (left) and from predictions of the validation cohort. **C** corresponding confusion matrices. (OSA: ODI > 30/h or ODI > 10/h & ESS > 10 points; control: ODI < 5/h or ODI < 10/h & ESS < 11 points; stratification 1)

## 4. Discussion

To the best of our knowledge, this is the first report of a validation of breath biomarkers for OSA. Previous studies using e-noses, offline gas-chromatography coupled to mass spectrometry, or enzyme immunoassays to analyze exhaled breath condensate have achieved promising results regarding the distinction between OSA patients and controls without OSA from exhaled breath.[19] However, sample sizes in all these studies were limited and none of the results has been validated in an independent cohort of patients. In this study, we could confirm in a large and independent cohort that breath intensities of many of our previously discovered potential biomarkers for OSA differ significantly between OSA patients and controls without OSA. Most of the markers are consistently increased in OSA patients. We could also confirm a correlation between breath signal intensity and disease severity (represented by the ODI) for several metabolites, supporting the association of these metabolites with apnea-related nocturnal hypoxemia. Our findings of correlations between breath signals and ESS scores indicate that not only hypoxia but also sleepiness is reflected in the metabolic breath pattern. The results from this study suggest that the 33 metabolites shown in Table 2 represent a panel of biomarkers, being robust enough concerning inter-individual variability to form a promising diagnostic tool. Inter-individual differences are the most likely reason for lower correlation coefficients between signal intensities in breath and ODI that we observed in this diverse validation cohort compared to the correlations between the signal intensities and the within-subject change of ODI upon CPAP withdrawal, which we reported previously.

**Table 2:**
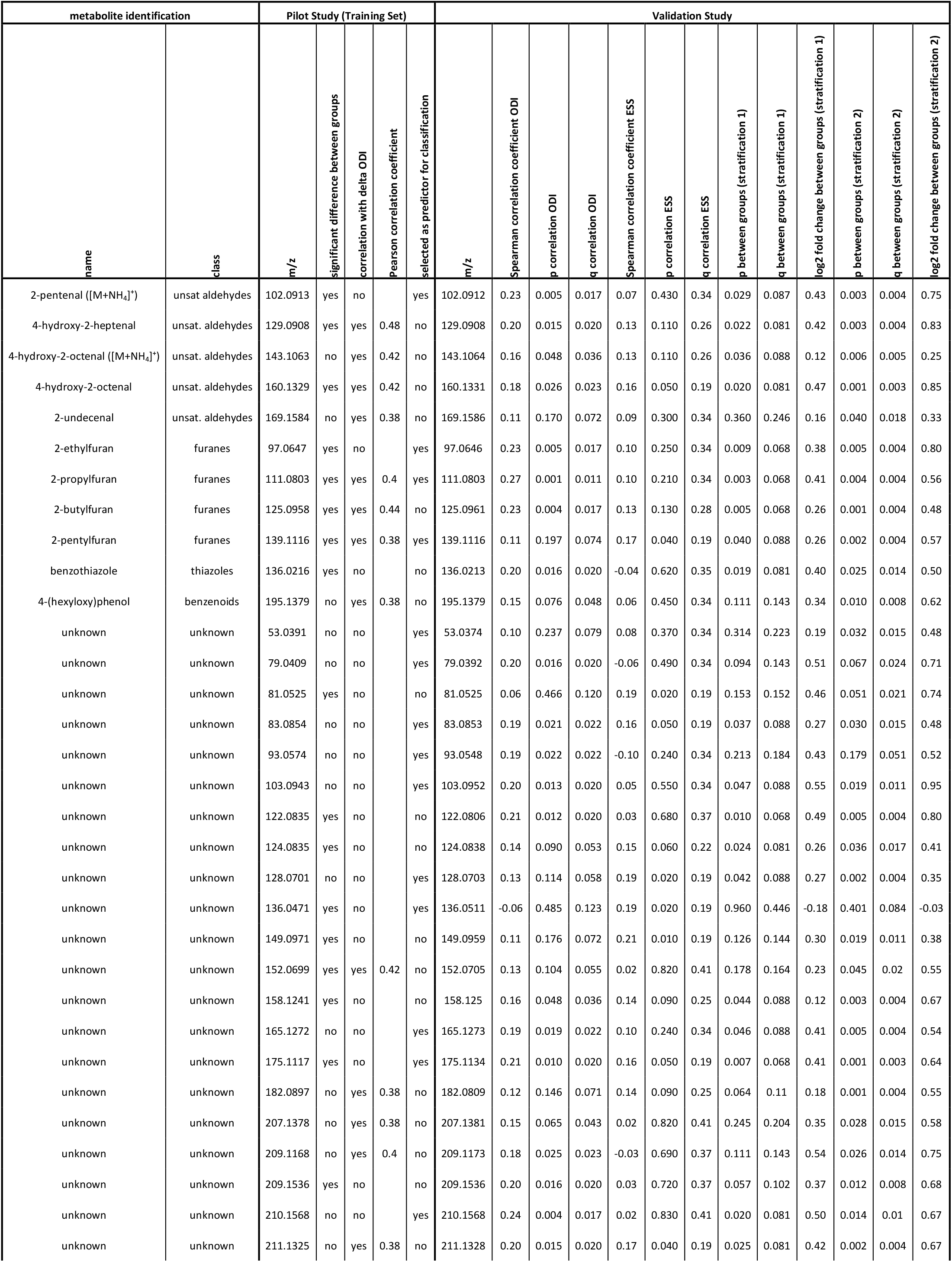
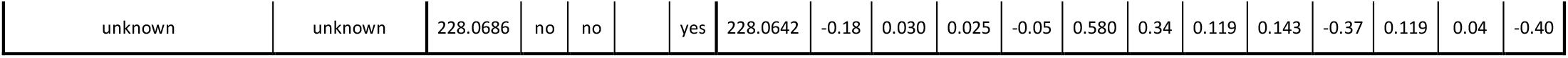
A panel of validated OSA biomarkers. 33 of the previously detected[24] biomarkers for OSA show a significant correlation with the ODI or ESS or significant differences between OSA patients and control subjects in the validation cohort (significance level: p < 0.05). (unsat. aldehydes: unsaturated aldehydes)

It seems unlikely that there is one single biomarker, which is sufficient for diagnosing a disease like OSA, associated with complex metabolic and cardiovascular consequences. In contrast, a pattern of several biomarkers is more likely to be disease-specific. Therefore, classification algorithms based on machine learning are convenient tools for making clinical diagnoses based on biomarker patterns. Here, we achieved a classification of the validation data set with an AUROC of 0.66, 76% sensitivity and 42% specificity, when we trained the model with the data from the independent patient cohort of our previously reported study.

Since the patient cohort of this study was much more diverse compared to the one in our pilot study, a lower classification performance would be expected. However, the support vector machine (SVM) model we used, performed already worse in the cross validation with the training data (AUC_CV_ = 0.59) compared to the model presented in the pilot study. This is likely to be due to different stratification criteria used in both studies, since with such small training sets, few samples can have a considerable influence. To test this hypothesis, we applied different stratification criteria. When we stratified the training data only based on ODI (as it was done in the pilot study), the model performance becomes comparable with the results that were reported previously (AUC_CV_ = 0.79, Figure A8A-C). This result supports our choice of an SVM model. However, the prediction of the validation data, which was also stratified only based on the ODI, not only failed to improve, but even declined slightly (AUC = 0.62, Figure A8A-C). This indicates that groups that are defined based on ODI and ESS can be distinguished better from the breath pattern with this biomarker panel than groups that are defined by ODI only. This is desirable, since a combination of ODI and ESS better reflects the clinical picture of significant OSA than ODI on its own. All performance measures of this classification procedure (classification 2) are reported in the supplementary results and are summarized in Table A2.

SESI is still a rather novel technique and its performance is constantly improving. Furthermore, in the time between the pilot study and this validation study, technical improvements have been implemented. For example, a new, more robust ion source[31], has been developed, and we adapted collision gas settings of the MS in order to prevent fragmentation, although this leads to increased cluster formation. This is most likely the reason why we did no longer detect all of the markers that were reported previously. The comparison of our data before and after batch correction between the data sets from both studies indicated that the data for the 78 potential biomarkers detected in both data sets are comparable and there is only a very small batch effect. A negligible change of the results from the classification procedure applied after batch correction confirmed this observation (classification 3, PCA plot and classification outcome are shown in Figure A2 and Figure A9, and classification results are described in supplementary results and Table A2).

Another factor that might impair the classification results is a lack of standardization of SESI-HRMS and the lack of real-time breath quality control samples, respectively, at the time when the study was conducted. Instrumental drifts are a common issue in large-scale metabolomics studies, which is overcome in offline-techniques with quality controls.[32] These samples are then used for normalization, i.e., to separate the biological variation of interest from unwanted technical variation or other confounding factors, such as exogenous influences. Since such samples are not yet available for real-time breath analysis, a higher degree of standardization of sampling and methodology is required.[33] In future studies, a reference gas mixture could be used to check the instrument performance and thereby reduce technical noise. This might improve the effectiveness of SESI-HRMS for screening for OSA or other metabolic conditions. Ideally, in a next step, calibration with standards of validated and identified biomarkers, such as the ones identified in this study, could be applied using standard addition. The standard addition procedure brings the advantage that in addition to technical fluctuations, matrix effects such as the influence of humidity on ionization efficiency, and ion suppression effects could be eliminated. However, the biggest challenge here is the availability of gaseous standards as well as the bottleneck of identification of metabolites. One possible approach is the use of permeation tubes.[34] Another confounding factor that might compromise the results are isobaric compounds since no separation step, such as chromatography, is used in real-time SESI-HRMS. To overcome this challenge, a targeted detection of the validated and identified biomarkers could involve MS-MS quantification or a coupling to ion mobility spectrometry and thereby provide a higher molecular specificity.

To date, different scores are derived from questionnaires for an initial approach to OSA screening. Our classification performance is comparable with the performances of the STOP-bang[14] and Berlin scores[15]. The NoSAS score performs slightly better, AUROCs of 0.74 and 0.81 have been reported from two different patient cohorts. However, in terms of sensitivity our results are comparable with the NoSAS score.[16] For screening, this is most relevant, since ideally no subjects with OSA are missed. It has been previously shown that the combination of NoSAS and metabolomics data can improve predictive performance remarkably.[35] Here, real-time breath analysis could speed up diagnosis and make it even less bothersome for the patients. We think that the combination of exhaled breath analysis and the NoSAS score might provide an objective and easy-to-perform assay for screening patients with possible OSA. Only positively tested patients would then need to undergo the time-consuming, costly and inconvenient respiratory polygraphy to confirm or refute the screening result. During screening, even multiple testing would be possible, since the breath test is fast and non-invasive. This might help to overcome the problem of a considerable night-to-night variation of OSA. Further studies, looking at the combination of the NoSAS score and SESI-HRMS are needed.

In addition, exhaled breath metabolomics can provide detailed information about metabolic changes because biological compounds can be identified. Here, we could confirm the association of unsaturated aldehydes, furans and benzothiazole with OSA. These findings are strengthening the hypotheses from our pilot study of increased oxidative stress levels and altered gut microbiota in OSA patients.[24] Moreover, our findings of metabolites such as 2-pentylfuran and 4-hydroxy-2-octenal, correlating with both, ESS and ODI, suggest an association of those metabolites with the sleep deprivation going along with OSA leading to increased sleepiness.

## 5. Conclusions

In conclusion, we could confirm our previous findings of an OSA specific metabolic breath pattern and validate a panel of 33 biomarkers in a larger and broader cohort of patients with possible OSA. This is the first validation study for breath analysis by SESI-HRMS, bringing this technique an important step closer to its application in clinics. However, being implemented for clinical use, the added value of SESI-HRMS measurements to conventional OSA screening questionnaires, such as NoSAS, should be evaluated in further patient cohorts.

## Data Availability

The original data and code used in this publication are made available in a curated data archive at ETH Zurich (https://www.research-collection.ethz.ch) under the DOI 10.3929/ethz-b-000456155.

https://www.research-collection.ethz.ch/handle/20.500.11850/456155

## Acknowledgements

We would like to thank Thomas Gaisl, Martin Gaugg, Marloes Maathuis, Sanzio Monti, Martin Osswald and Bettina Streckenbach for valuable scientific discussion. This project is part of the Zurich Exhalomics project, a flagship project of University Medicine Zurich.

