## Supplementary Information for "Validation of breath biomarkers for obstructive sleep apnea"

### Appendix A. Supplementary Information

**Supplementary results:**

The classification performance that we estimated with this stratification in the cross-validation based on the training data is considerably lower than the one reported in the pilot study (reported $AUC_{cross\text{-}validation} = 0.87$). Previously, only the ODI was used for stratification. To evaluate whether the metabolic pattern of exhaled breath predicts groups defined only by ODI better, we repeated the classification procedure again with the following grouping criteria: ODI > 30/h (OSA), ODI < 10/h (control) (Figure A8d). For the training set we reduced the control group to ODI < 2/h in order to get balanced group sizes (Figure A8). With these stratification criteria we achieved similar predictive performance in the training set as in the pilot study ($AUC_{cross\text{-}validation} = 0.79$, Figure A8b and c). Nevertheless, the prediction of the validation set declined to AUC = 0.62 (Figure A8e and f).

Since technical improvements have been made between the pilot study and this validation study and both studies were time-wise separated, we assessed the comparability of both data sets in a principal component analysis (Figure A2a). Even though only a slight shift between the two sets is noticeable, we corrected it successfully by applying a batch correction algorithm based on an Empirical Bayes method (Figure A2b). We subsequently repeated the classification procedure with the adjusted data. The results are presented in Figure A2. However, the AUC improved only by 0.01 to 0.67. The results from all classification procedures are summarized in Table A2.

**Supplementary discussion:**

We further investigated the samples that were false negatives in all three classification procedures as indicated in Figure A10. One of them was wearing lipstick, which we observed often to be confounding due to ion suppression caused by prominent plasticizers or other ingredients with a good ionization efficiency. Even though visual inspection of the spectra of this study participant did not capture our attention, it is possible that a contamination from the lipstick compromised the results. The three remaining false negatives have in common that the respiratory polygraphy was not carried out on the same day as the SESI-HRMS breath measurement. It could thus be, that the MS-based diagnosis does not match the clinical diagnosis due to night-to-night variability of OSA, which is well-known. To overcome this problem, multiple measurements would be required.

**Supplementary Figures:**

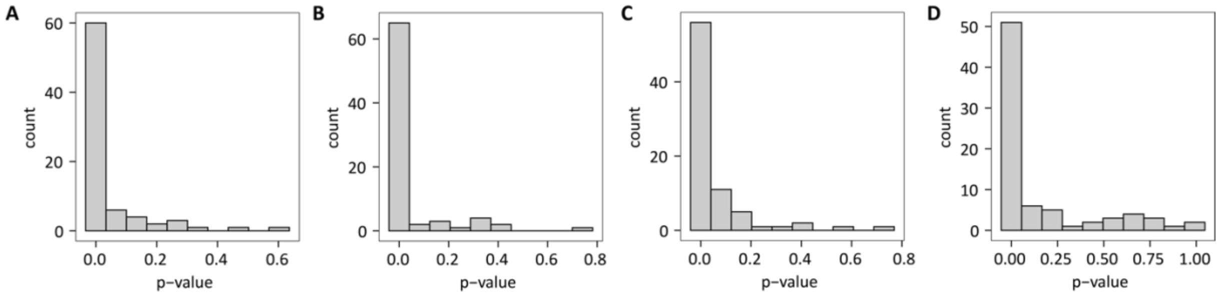


**Figure A1:** P-value distributions from Shapiro-Wilk's test for normality. **A** Control group from stratification 1 (OSA: ODI > 30/h or ODI > 10/h & ESS > 10 points, control: ODI < 5/h or ODI < 10/h & ESS < 11 points). **B** OSA group from stratification 1. **C** Control group from stratification 2 (control: ODI < 10/h & ESS <11 points, OSA: ODI < 30/h & ESS > 10 points). **D** OSA group from stratification 2.

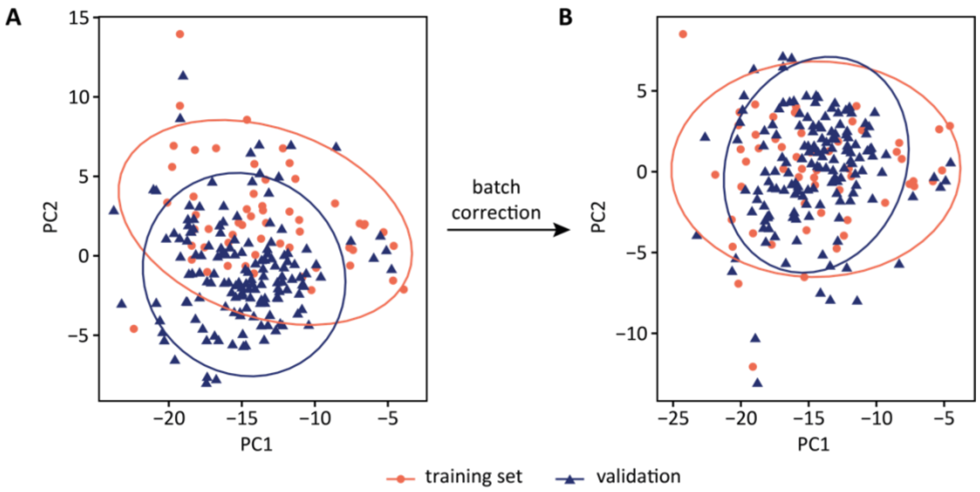


**Figure A2:** PCA plot before (**A**) and after (**B**) batch correction.

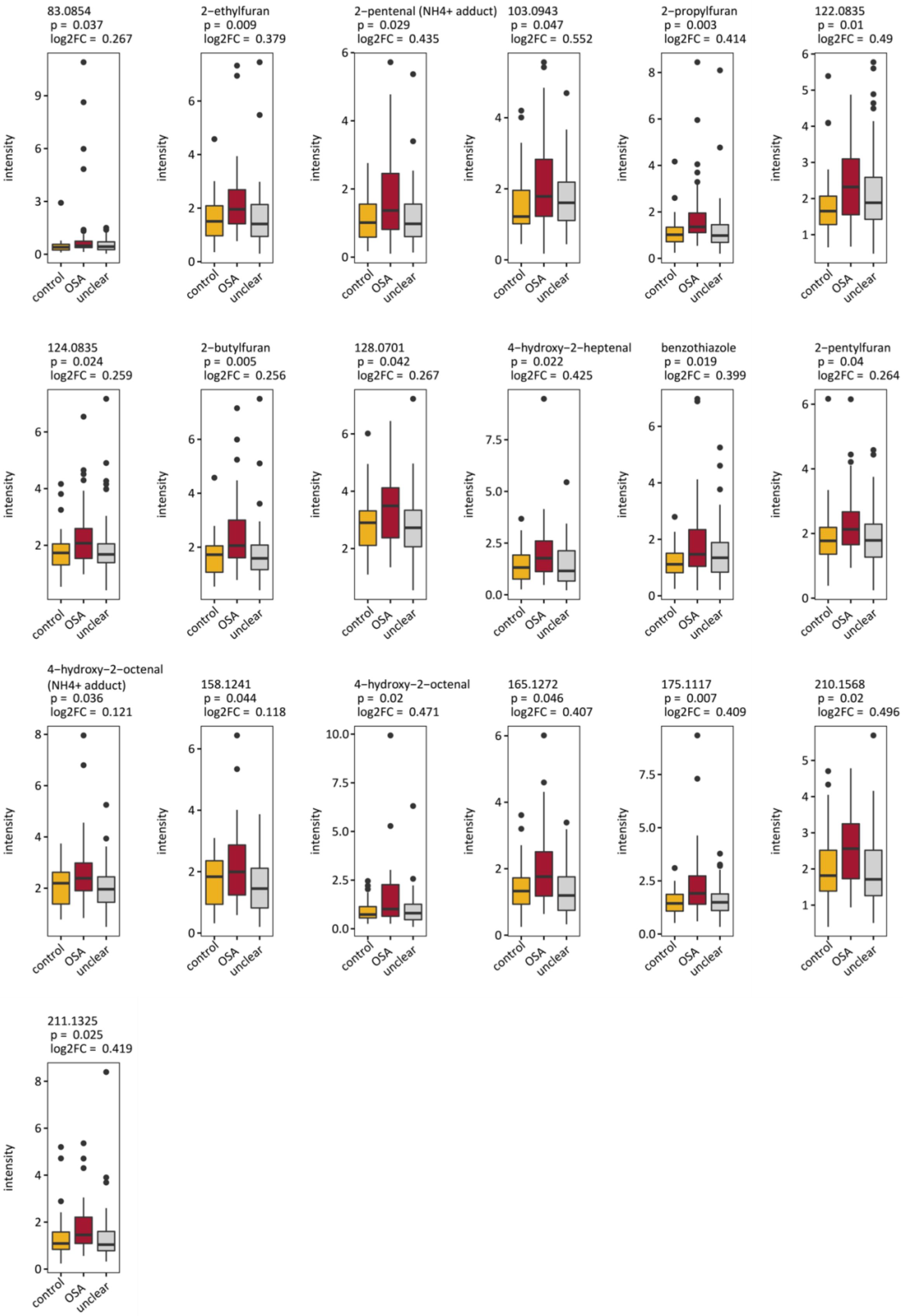


**Figure A3:** Boxplots (center line: median, box limits: 25th and 75th percent quantile, whisker length: 1.5 interquartile range) for all features with significant differences between OSA patients and subjects without OSA. (OSA: ODI > 30/h or ODI > 10/h & ESS > 10 points; control: ODI < 5/h or ODI < 10/h & ESS < 11 points; unclear: in between; stratification 1). log2FC: log2 fold change, p: p-value.

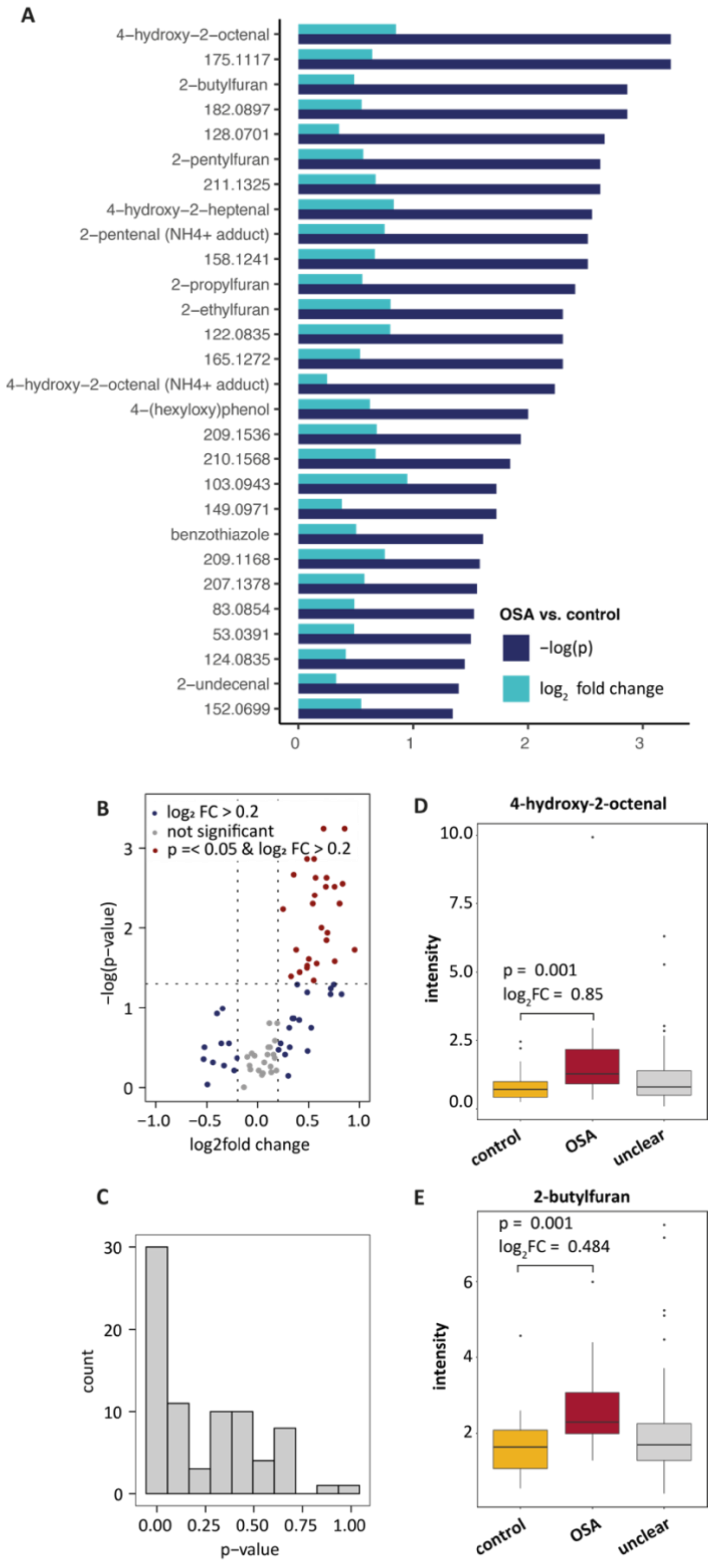


**Figure A4:** Significant differences in metabolic breath patterns between OSA patients and individuals without OSA (control: ODI < 10/h & ESS <11 points; OSA: ODI < 30/h & ESS > 10 points; unclear: in between; stratification 2). **A** p-values and fold changes of significant features sorted by significance. **B** volcano plot for all 78 metabolites. **C** p-value distribution for between-group differences from Mann-Whitney-U test. **D**, **E** Exemplary boxplots (center line: median, box limits: 25th and 75th percent quantile, whisker length: 1.5 interquartile range) of 4-hydroxy-2-octenal and 2-butylfuran. Boxplots for all significant features are provided in Figure A3, numeric results for significant features are summarized in Table 2 and numeric results of all 78 features are given in Table A1.

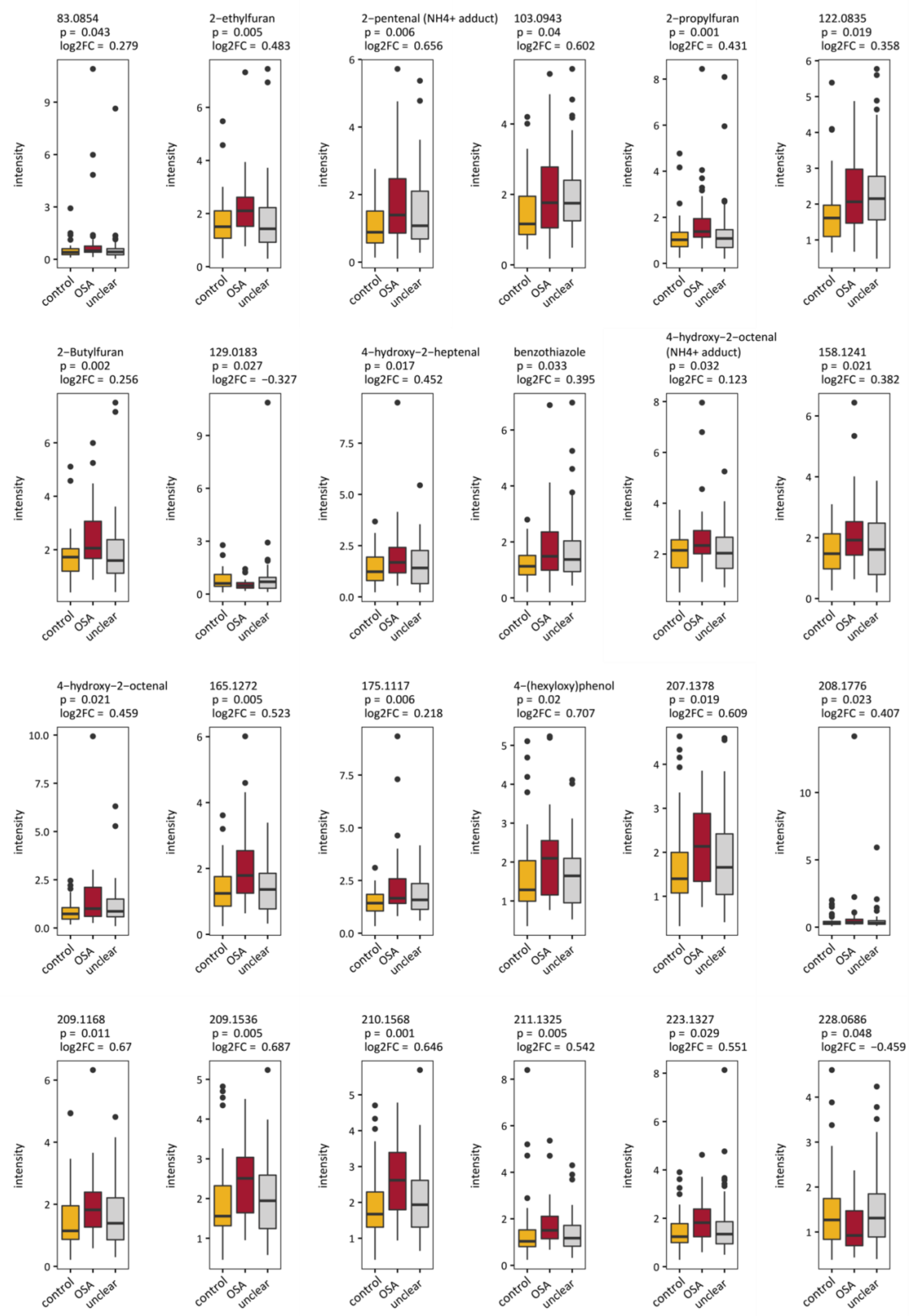


**Figure A5:** Boxplots (center line: median, box limits: 25th and 75th percent quantile, whisker length: 1.5 interquartile range) for all features with significant differences between OSA patients and individuals without OSA (control: ODI < 10/h & ESS <11 points; OSA: ODI < 30/h & ESS > 10 points; unclear: in between; stratification 2).

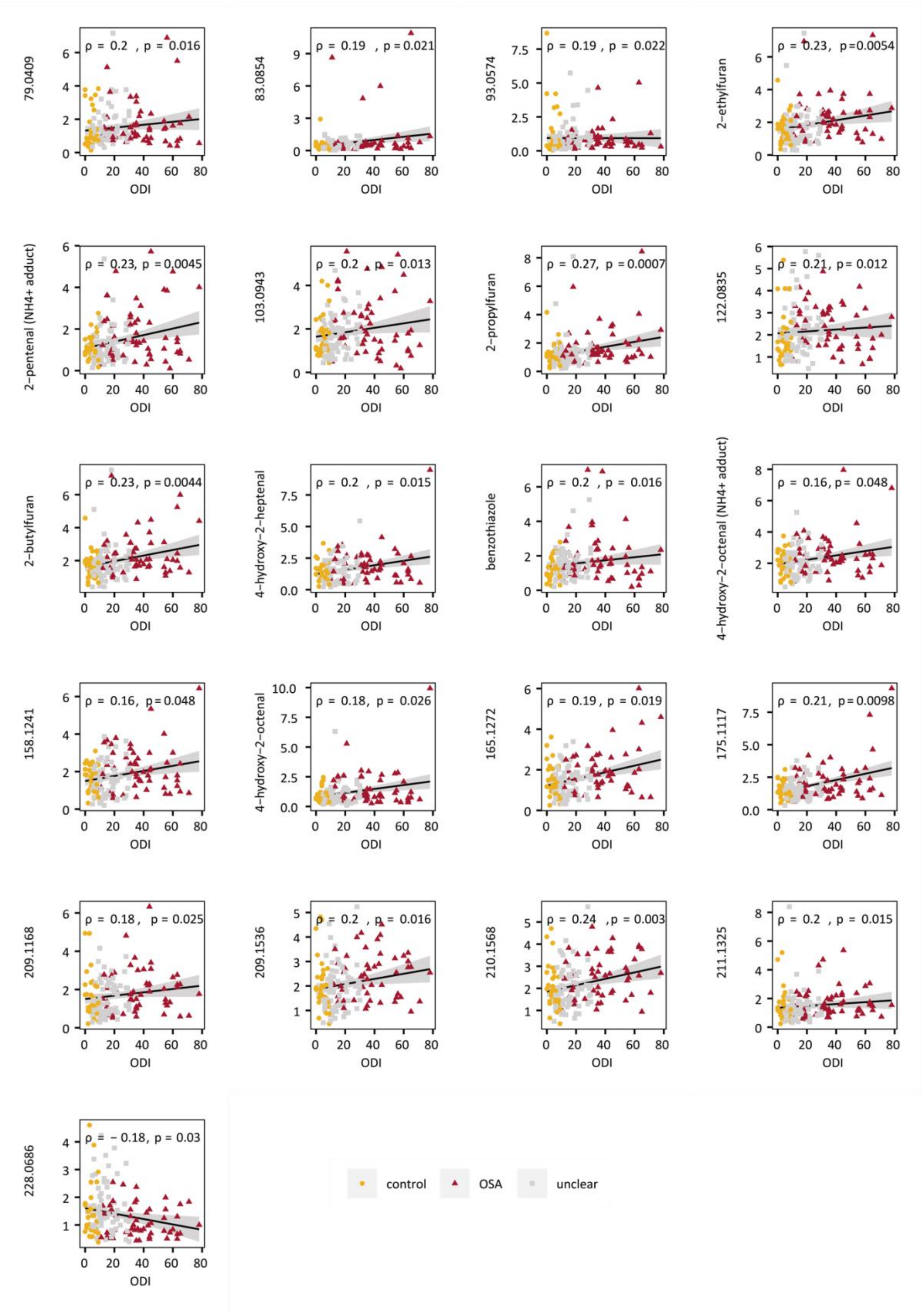


**Figure A6:** Linear regression for all features that correlate significantly ($p \leq 0.05$) with ODI (OSA: ODI > 30/h or ODI > 10/h & ESS > 10 points; control: ODI < 5/h or ODI < 10/h & ESS < 11 points; unclear: in between; stratification 1).

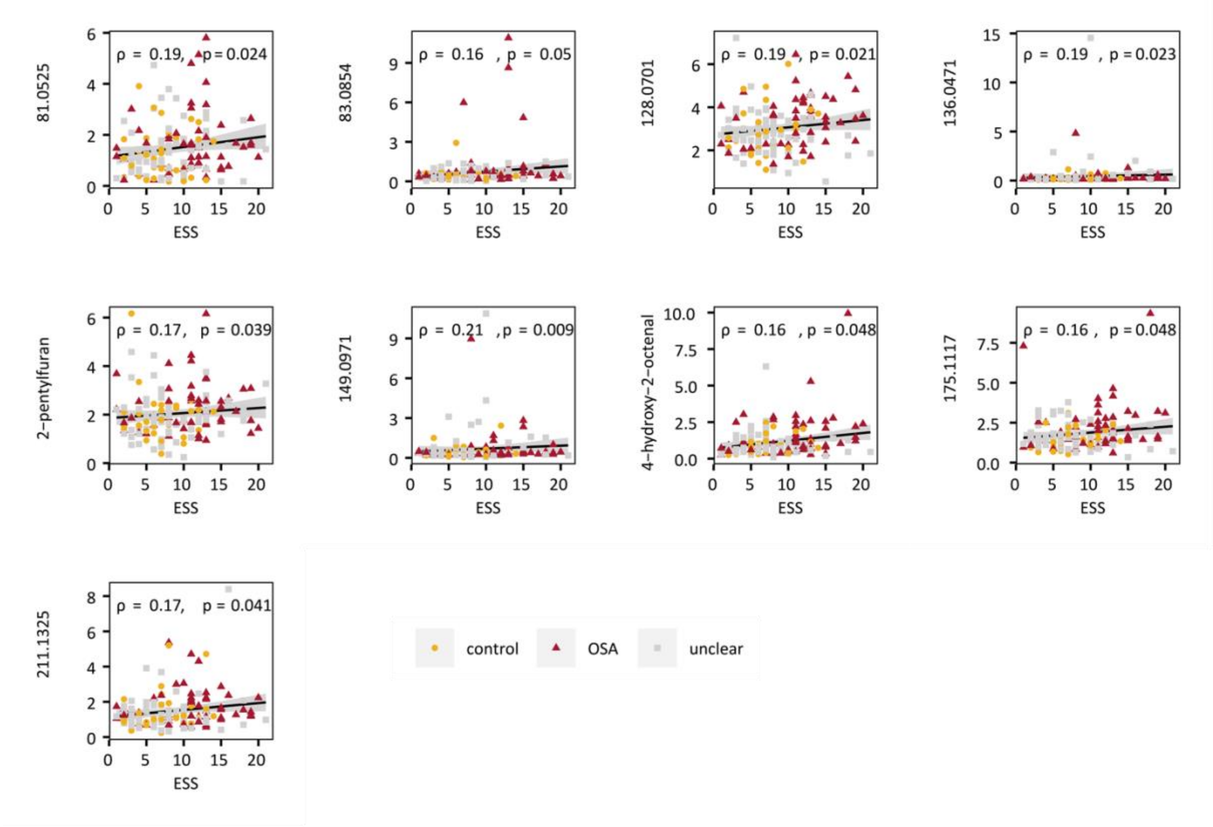


**Figure A7:** Linear regression for all features that correlate significantly ($p \leq 0.05$) with ESS (OSA: ODI > 30/h or ODI > 10/h & ESS > 10 points; control: ODI < 5/h or ODI < 10/h & ESS < 11 points; unclear: in between; stratification 1).

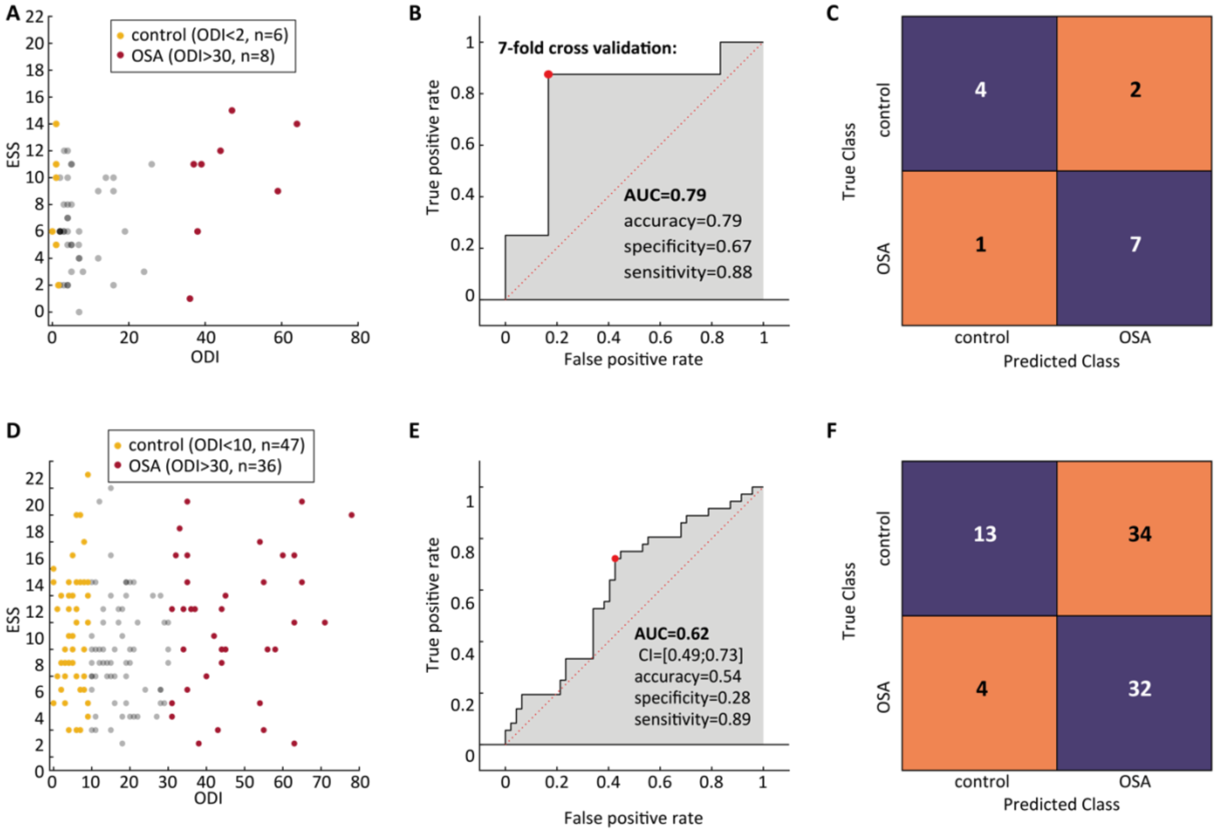


**Figure A8:** Classification results with stratification similar to pilot study (classification 2). **A** ESS and ODI of samples in the training set. **B** ROC curve from 7-fold cross validation of the classification model with the training set and **C** corresponding confusion matrix. **D** ESS and ODI of the samples in the validation cohort. **E** ROC curve from predictions of the validation cohort and **F** corresponding confusion matrix.

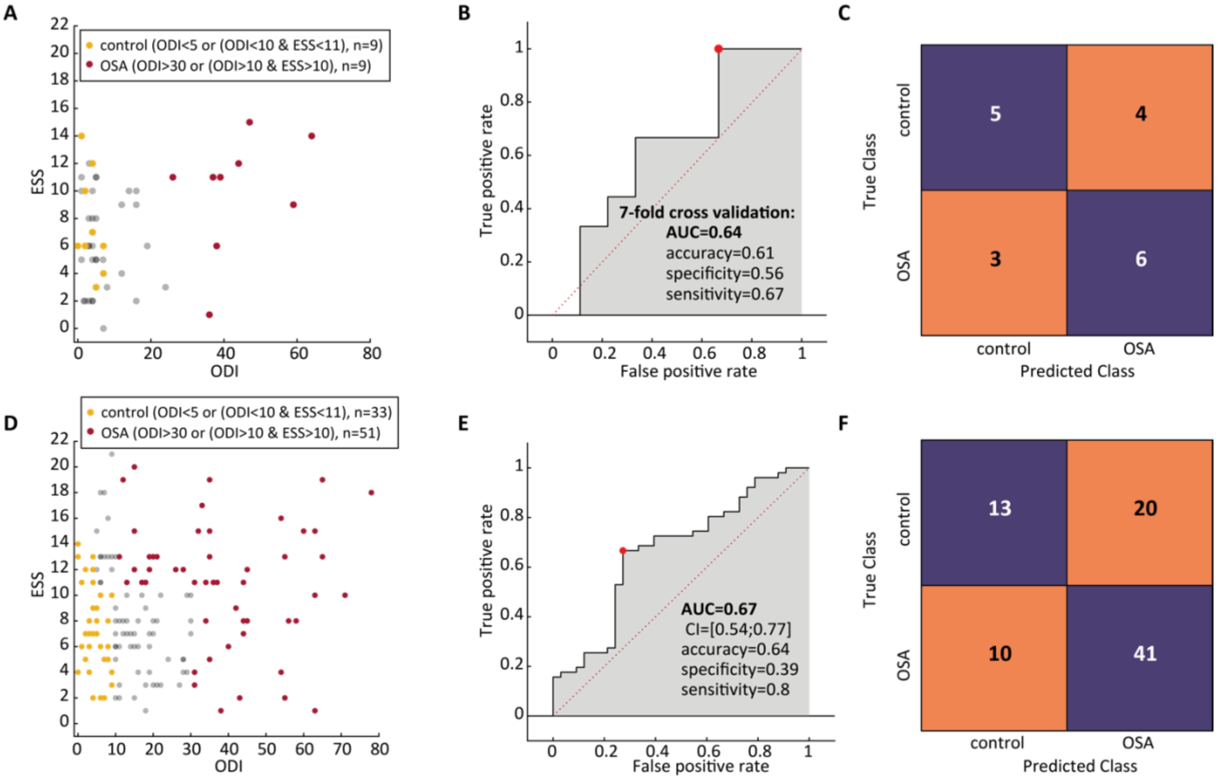


**Figure A9:** Classification results after batch correction (classification 3). **A** ESS and ODI of samples in the training set. **B** ROC curve from 7-fold cross validation of the classification model with the training set and **C** corresponding confusion matrix. **D** ESS and ODI of the samples in the validation cohort. **E** ROC curve from predictions of the validation cohort and **F** corresponding confusion matrix.

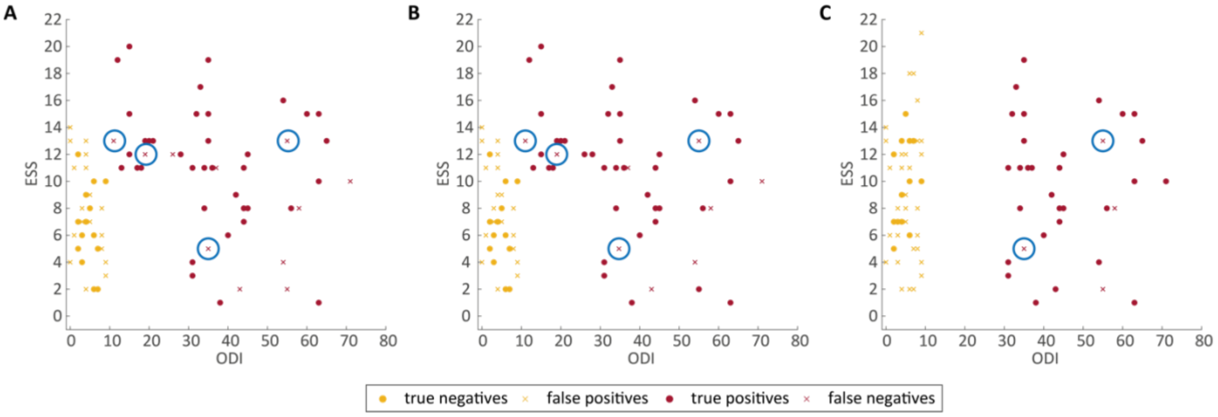


**Figure A10:** Accuracy of diagnosis based on metabolic pattern of exhaled breath. Correctly and wrongly predicted samples of validation data set in classification 1 **(A)**, in classification 2 **(B),** and in classification 3 **(C)**. Samples that are false negatives in all three classification procedures are highlighted with blue circles.

**Supplementary Tables:**

**Table A1:** Results from statistical analysis of validation data for all 78 features that have been reported previously as potential biomarkers of OSA. (see separate spreadsheet)

**Table A2:** Diagnostic performance of exhaled breath analysis for OSA. (AUC: area under receiver operating characteristic curve; CV: cross validation; TPR: true positive rate, FPR: false positive rate, TNR: true negative rate, FNR: false negative rate, CI: confidence interval)

| | Training Set | | | | | | | | Validation Set | | | | | | | | | |
|---|---|---|---|---|---|---|---|---|---|---|---|---|---|---|---|---|---|---|
| | **stratification** | **$n_{control}$** | **$n_{OSA}$** | **AUC(CV)** | **TPR** | **FPR** | **TNR** | **FNR** | **stratification** | **$n_{control}$** | **$n_{OSA}$** | **AUC** | **CI1** | **CI2** | **TPR** | **FPR** | **TNR** | **FNR** |
| **classification 1** | **OSA**: ODI>30 or (ODI>10 & ESS>10) **control:** ODI<5 or (ODI<10 & ESS<=10) | 9 | 9 | 0.59 | 67% | 33% | 67% | 33% | **OSA**: ODI>30 or (ODI>10 & ESS>10) **control**: ODI<5 or (ODI<10 & ESS<11) | 33 | 51 | **0.66** | 0.55 | 0.79 | 76% | 58% | 42% | 24% |
| **Classification 2** (stratification similar to pilot study) | **OSA**: ODI>30 **control**: ODI<2 | 6 | 8 | 0.79 | 88% | 33% | 67% | 13% | **OSA**: ODI>30 **control**: ODI<10 | 47 | 36 | **0.62** | 0.49 | 0.73 | 89% | 72% | 28% | 11% |
| **classification 3** (after batch correction) | **OSA**: ODI>30 or (ODI>10 & ESS>10) **control**: ODI<5 or (ODI<10 & ESS<=10) | 9 | 9 | 0.64 | 67% | 44% | 56% | 33% | **OSA**: ODI>30 or (ODI>10 & ESS>10) **control**: ODI<5 or (ODI<10 & ESS<11) | 33 | 51 | **0.67** | 0.54 | 0.77 | 80% | 61% | 39% | 20% |
